# Modern approaches to predicting vaccine hesitancy: A scoping review

**DOI:** 10.1101/2025.01.29.25321367

**Authors:** Keshav Gandhi, Sami Alahmadi, Rosie Hanneke, Alexander Gutfraind

## Abstract

**Introduction:** Motivated by the disproportionate burden of infectious diseases on vulnerable populations and the risk of future pandemics, we conducted a scoping review to analyze the state of the literature about “vaccine uptake indices,” defined as models that predict vaccination rates by geospatial area. We analyzed novel vaccine uptake indices created in response to the recent COVID-19 pandemic. The aim of this scoping review is to survey the state of the literature regarding vaccine uptake indices relating to COVID-19 and other infectious diseases.

**Methods:** We conducted a scoping review with a systematic search strategy to identify relevant articles from the databases Embase, PubMed, and Web of Science with title and abstract screening, full-text review, and data extraction.

**Results:** Database searches resulted in 3,615 potential articles, of which 229 reports were included. Fifteen studies (7%) were determined to be methodologically advanced vaccine uptake indices that had at least three of the following characteristics: the use of individual- and population-level predictor variables (100 [44%]), geo-spatiotemporal analysis (58 [25%]), data usage agnostic to vaccine specificity (50 [22%]), or sociobehavioral frameworks of health (such as the Health Belief Model and Theory of Planned Behavior) (30 [13%]).

**Conclusion:** This scoping review offers suggestions for future research of next-generation vaccine uptake indices before use in vaccination campaigns of recurring or novel infectious diseases. Areas to pursue include utilizing individual-level data about vaccination behaviors in conjunction with administrative data, solving the challenge of implementing small-area spatiotemporal analysis, using vaccine-agnostic methods that consider data from more than one infectious disease, and assisting causal inference with theoretical frameworks.

## 1. Introduction

Even for highly effective and safe vaccines, vaccination rates range from approximately 50% to 90% in the United States [1], [2], and there are concerns about disparities in vaccination rates across different geographic units and populations [3], [4]. To help target resources to address these inequities, many studies constructed vaccine uptake indices (VUIs), defined as quantitative models that predict a vaccination rate within a given geographic area [5], [6], [7]. Such indices can address the need for equitable, targeted vaccination campaigns, which has been exemplified through the recent COVID-19 pandemic [4], [8], [9].

Vaccine uptake is closely related to the concepts of vaccine acceptance and vaccine hesitancy [10]. Vaccine uptake can be time-dependent [11], especially when a vaccine is in short supply, but at any given time point, there are often persistent inequalities in vaccination rates by group and geography [12], [13], [14], [15]. While the literature outside of COVID-19 is less advanced, development of VUIs could prove useful in the case of vaccination for other infectious diseases, such as human immunodeficiency virus (HIV), hepatitis C virus (HCV), or Epstein-Barr virus (EBV), where vaccine development continues to be of significant interest [16], [17], [18]. These tools could employ data on factors such as social stigma, perceived risk, and access to healthcare to help predict potential vaccine acceptance in a given area. Similarly, predicting vaccine acceptance may be valuable for vaccines that (1) are currently in early stages of development or clinical trials, including the HCV and EBV vaccines, (2) have been newly developed and distributed, such as the respiratory syncytial virus (RSV) vaccine [19], or (3) are mature vaccines with insufficient uptake, such as the influenza or human papillomavirus (HPV) vaccines [20]. Prediction of vaccination for both existing (e.g., influenza) and novel infectious diseases remains important and challenging [21], [22], [23]. Identifying regions likely to have increased vaccine hesitancy allows for targeted preventive strategies aimed at addressing barriers to acceptance. However, in current practice, VUIs are not used on a wide scale to predict vaccine acceptance in populations, and as a result, many studies utilize vulnerability indices to predict vaccine uptake and influence policy [24], [25].

A vulnerability index is a quantitative model that predicts the impact of an infection (e.g., COVID-19 infections or mortality) by area based on geospatial area and social risk, allowing for further analysis of inequities [26], [27]. Some vulnerability indices may be used to predict vaccine uptake outside of their intended applications. For instance, the Center for Disease Control’s Social Vulnerability Index (SVI) can be used to predict vaccination levels, but it was designed to assess the impact of natural disasters on a population by geospatial area [28]. It includes factors—such as the proportion of children, single-parent households, and households with access to a personal vehicle—that increase vulnerability during evacuation but likely do not significantly affect infectious disease risks [29]. Its updated form, the COVID-19 Community Vulnerability Index (CCVI), takes public health infrastructure into account but was designed to predict COVID-19 vulnerability trends and overall community risk rather than vaccination uptake by small-area analysis [30]. Because many vulnerability indices are not specifically designed to predict vaccination rates, introducing VUIs may facilitate equitable resource allocation with increased accuracy [8], [31], [32].

In this review, we survey the state of the literature regarding VUIs using the case of COVID-19. To our knowledge, this is the first study of its kind in the literature. Other meta-analyses and systematic reviews focus on either aggregating data on factors that contribute to COVID-19 vaccination inequities or creating a pooled vaccine acceptance rate from individual studies [33], [34], [35].

The overall goal of this study is to support the development of next-generation VUIs that address vaccination disparities by focusing on key areas identified in the current literature. We aim to determine the scope of the existing literature that explores quantitative indices that predict vaccination outcomes and/or social vulnerability. We evaluate whether the current literature: (1) makes use of quantitative models that predict vaccine uptake and social risk by geospatial area, (2) combines different sources of predictive variables or features of VUIs, including individual-level predictor variables [36], [37], geo-spatiotemporal analysis [38], [39], and vaccine agnosticity [21], [22], and (3) uses sociobehavioral frameworks to limit model misspecification [40], [41].

Due to limited data availability and the need for promptness of research during the COVID-19 pandemic, we hypothesized that most geospatial VUIs would directly predict vaccination using solely administrative data (i.e., vaccine uptake records from public health agencies), with limited use of models trained using data from infectious diseases beyond COVID-19 or frameworks to assist causal inference.

### 2. Methods

To execute the review, we used Arksey and O’Malley’s scoping review framework and limited the search to VUIs that have been newly developed in response to the COVID-19 pandemic [42]. We chose to conduct a scoping review because it is a flexible methodology that is inclusive of heterogeneous study designs and interdisciplinary areas of interest. This scoping review is reported according to the PRISMA-ScR reporting guidelines [43].

A list of key words and phrases was constructed to retrieve papers relevant to VUIs from the literature. Inclusion and exclusion criteria were developed in a review process based on an initial exploratory survey of the scope of literature. We then found potential exemplar vaccine uptake and/or novel vulnerability indices and reframed our search strategy to include such articles [8], [24], [25], [26], [38]. Keywords and controlled vocabulary (Medical Subject Headings and Emtree terms) were grouped into four conceptual domains separated by “AND” operators: Quantitative Indices, Geospatial Analysis, Infectious Disease (COVID-19), and Vaccination or Vulnerability (Outcomes). Within each domain, search terms were separated by “OR” operators. MeSH terms in PubMed (and analogous controlled vocabularies in other databases) were used to generate additional search terms within each domain (see Supplement 1).

Searches were limited to publications from November 1, 2019 to the date of database retrieval in order to exclude articles not related to COVID-19. The inclusion criterion was to find studies that applied existing or novel indices to predict COVID-19 vaccination rates or social vulnerability, and the exclusion criterion was the lack of prediction of vaccine uptake or hesitancy (VUIs) or infection or mortality rates (vulnerability indices).

On November 7, 2022, we searched the database Web of Science (including Science Citation Index Expanded, Social Sciences Citation Index, Arts & Humanities Citation Index, and Emerging Sources Citation Index). On November 9, 2022, we searched the databases Embase.com and PubMed (including MEDLINE).

The search results were uploaded to citation management software [44], where duplicate records were removed before title and abstract screening. Two independent reviewers blindly completed title and abstract screening and resolved decision conflicts through discussion in order to select studies to be reviewed in the full text. This process was repeated for the full-text review.

Data charting was performed with the included studies. Themes and areas of interest generated from the full-text review stage were compiled into an extraction form for reviewers to complete (see Supplement 2). This involved the creation of a coding system designed to classify different types of articles and operationalize definitions in order to better differentiate between classes of articles. After the coding system was agreed upon by the full-text reviewers, data extraction was performed to generate the dataset, and statistics were calculated using R [45].

## 3. Results

Database searches retrieved 7,156 records (Fig. 1). After removing duplicates, 3,615 records remained for screening. Following the title and abstract screening, 345 reports were assessed for eligibility. A total of 229 reports met the inclusion criteria and were included in the scoping review. See Supplement 3 for a list of further reading and Supplement 4 for each included report in Supplement 1.

**Fig. 1.**
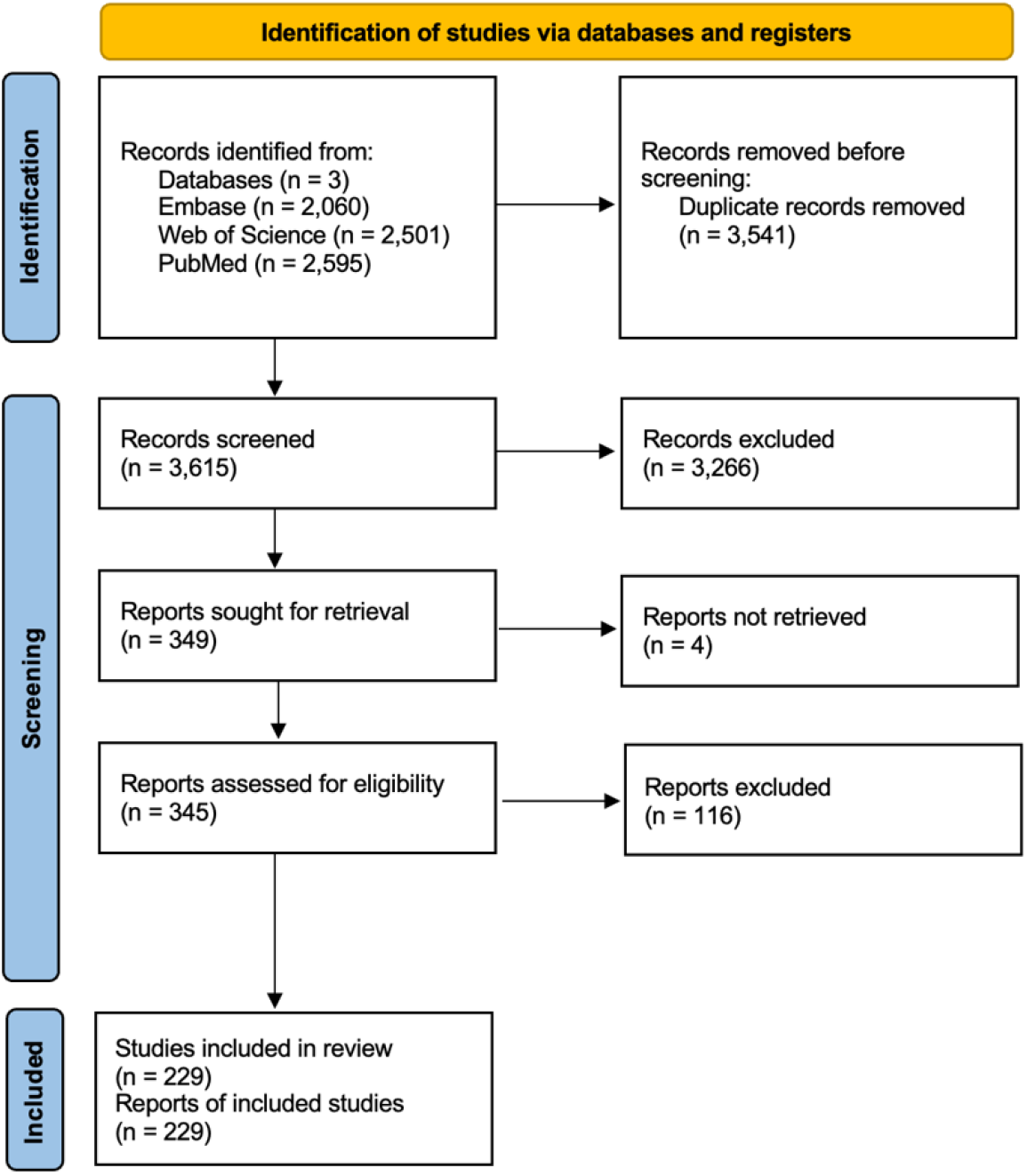
A PRISMA flow diagram depicting the scoping review process [46].

Of the 229 included studies, 203 (89%) were classified as VUIs and 26 (11%) were classified as vulnerability indices. The studies demonstrated a variety of methodological approaches, as presented in Table 1. Specifically, 151 VUIs (66%) utilized geospatial data, and of this group, 58 (38%) incorporated geo-spatiotemporal analysis. A total of 50 studies (22%) integrated data from multiple infectious diseases in their analyses, and 30 studies (13%) employed social and behavioral frameworks. Overall, 15 reports (7%) were classified as methodologically advanced VUIs, based on usage of at least three of the four following characteristics: a combination of individual- and population-level predictor variables, spatiotemporal analysis, vaccine-agnostic data usage, and health-related sociobehavioral frameworks.

**Table 1.** Vaccine uptake index characteristics by study design.

| Characteristic of Analysis<sup>a</sup> | All Studies<br>(n = 229) | Vaccine Uptake<br>Indices Only<br>(n = 203) | Vulnerability<br>Indices Only<br>(n = 26) |
| --- | --- | --- | --- |
| Aggregate-level data only | 105 (46%) | 79 (39%) | **26 (100%)** |
| Both aggregate- and<br>individual-level data<sup>b</sup> | 100 (44%) | 100 (49%) | 0 (0%) |
| Geospatial analysis | 151 (66%) | 125 (62%) | **26 (100%)** |
| Geo-spatiotemporal analysis<sup>b</sup> | 58 (25%) | **49 (24%)** | **9 (35%)** |
| Vaccine-agnostic elements<sup>b</sup> | **50 (22%)** | 46 (23%) | 4 (15%) |
| Sociobehavioral framework<sup>b</sup> | 30 (13%) | **30 (15%)** | 0 (0%) |
| Two characteristics<br>of interest | 52 (23%) | 51 (25%) | 1 (4%) |
| Three characteristics<br>of interest<sup>c</sup> | 15 (7%) | **15 (7%)** | 0 (0%) |
| Four (all) characteristics<br>of interest | 0 (0%) | 0 (0%) | 0 (0%) |

<sup>a</sup>Important trends are bolded<sup>b</sup>Characteristic of a methodologically advanced vaccine uptake index<sup>c</sup>Coded as a methodologically advanced vaccine uptake index

The studies used different geospatial levels for their analyses (Table 2). Small-area analysis was the most commonly used, including county- or city-level data (64 [42%]) and ZIP-code- or neighborhood-level data (33 [22%]). Among the studies that applied social frameworks, the Health Belief Model was the most commonly used (16 [53%]). Other notable frameworks included the Theory of Planned Behavior (8 [27%]) and the Protection Motivation Theory (2 [7%]). Four studies (7%) used more than one of these frameworks to analyze vaccination uptake.

**Table 2.** Categories of included geospatial levels and sociobehavioral frameworks.

| **Geospatial Levels** |  |
| --- | --- |
| Continent | 1 (1%) |
| Country | 19 (13%) |
| Country; state or province | 1 (1%) |
| State or province | 20 (13%) |
| State or province; county or city | 23 (15%) |
| County or city | 54 (36%) |
| County or city; ZIP code or neighborhood | 10 (7%) |
| ZIP code or neighborhood | 23 (15%) |
| **Sociobehavioral Frameworks** |  |
| Health Belief Model [47] | 12 (40%) |
| Health Belief Model, Theory of Planned Behavior | 2 (7%) |
| Health Belief Model, Theory of Planned Behavior, and others | 2 (7%) |
| Protection Motivation Theory [48] | 2 (7%) |
| Theory of Planned Behavior [49] | 4 (13%) |
| Other (used by one study only)<sup>a</sup> | 8 (27%) |

<sup>a</sup>Additional social and/or behavioral frameworks include the following: fear of adverse effects, knowledge, attitudes, and practices, moral foundations theory [50], prosocial priming, public fear of COVID-19, social identity approach, technology acceptance model [51], and theory of reasoned action [52]

### 3.1 Qualitative Trends

After the title and abstract screening and full-text review stages, two general types of study designs were primarily found by the study: Group A (cross-sectional surveys) and Group B (indices that make use of administrative data).

Group A studies, which comprised the plurality of the 203 VUIs, primarily focused on analyzing vaccine uptake within specific geospatial areas. These studies utilized individual-level variables, such as attitudes towards vaccination or comfortability with risk, to predict vaccine hesitancy, uptake, or outcomes. Multiple logistic regression was the most common analytical method employed (50 [25%]). Studies that used social or behavioral frameworks for model specification (30 [15%]) were included as Group A studies (Table 1).

Group B studies, although less frequently categorized as VUIs, focused on the use of aggregate-level variables, such as the association of vaccination rates with other public health outcome variables. Studies in this group used diverse methodologies to analyze administrative data, including random forests, Bayesian hierarchical modeling, and spatial regression [6], [8], [53]. Group B articles were also included as ecological studies or vulnerability indices (26 [11%]) and are represented in the “Vulnerability Indices Only” column of Table 1. All of the studies within this column (26 [100%]) used geospatial analysis and aggregate-level predictors of vaccine uptake without the use of individual-level predictors. Geo-spatiotemporal analysis was used at a higher proportion (9 [35%]) compared to vaccine uptake indices (49 [24%]).

## 4. Discussion

This systematic scoping review analyzed VUIs and vulnerability indices in response to COVID-19. Out of the 229 studies reviewed, 203 (89%) were categorized as VUIs and 26 (11%) as vulnerability indices. The review revealed a significant methodological diversity in the development of VUIs (Table 1). Specifically, 124 studies (54%) used any form of individual-level data regarding vaccination behaviors as predictor variables. One hundred twenty-five of the VUIs (62%) utilized geospatial analysis, but only 49 (24%) made use of geo-spatiotemporal analysis. Additionally, most geospatial analysis was performed at the county or city level (64 [42%]) (Table 2). Only 50 studies (22%) used data from multiple infectious disease sources, and 30 VUIs (15%) employed social and behavioral frameworks. Most of the studies that used a social or behavioral framework used the Health Belief Model (16 [53%]).

The VUIs demonstrated various approaches in their construction. The use of geospatial data was prevalent, with most analyses performed at the county or city level, suggesting the importance of county-level administrative data in small-area analysis. However, the limited use of data sources from more than one infectious disease in constructing these indices suggests a gap in leveraging patterns of historical vaccination behaviors to predict vaccination for novel pathogens. Moreover, the sparse application of social and behavioral frameworks points to an underutilized avenue for facilitating causal inference with VUIs. This review also highlights the potential for studies that could integrate individual-level predictor variables with currently used designs that utilize aggregate-level data.

The review identified two primary types of study designs: Group A (cross-sectional surveys) and Group B (indices using administrative data). Group A studies, which constituted the majority of VUIs, focused on analyzing vaccine uptake within specific geospatial areas using individual-level variables such as vaccination attitudes and risk comfortability. Group B studies more often utilize machine learning, nonparametric modeling, or other novel statistical approaches that may better capture complexity of nonlinear relationships between epidemiological variables, but Group A studies place a valuable emphasis on using individual-level variables as predictors. The usage of simple machine learning models by Group A studies, such as multiple logistic regression, increases the overall interpretability of these VUIs, but this may come at the expense of accuracy [54].

This individual-focused approach allows for the identification of specific behavioral and psychological factors that may inform targeted interventions, often through using theoretical frameworks of vaccine hesitancy to reduce model misspecification [55]. However, Group B studies are still important in understanding the impact of social vulnerability on vaccination rates. These tradeoffs are important to consider, and an ideal VUI may make use of both individual-level and population-level variables when making predictions across geospatial areas and avoiding the ecological fallacy.

A minority of study designs did not fit in either category. For instance, some of the studies constructed temporal dynamics models that utilized vaccination data from Google Trends, Twitter, Reddit, or other internet- or social media-based sources to make predictions about vaccination rate by geospatial area [36], [37], [56]. These types of studies uniquely combine data from individuals as well as administrative public health data sources and are also methodologically diverse in terms of their underlying statistical modeling techniques.

We have also found that the utility of VUIs often overlaps with that of vulnerability indices. In practice, both types of models are generally used to predict vaccination given a geospatial area [8], [25], [31]. Some ecological study designs compare the performance of multiple vulnerability indices that are currently used in the field (such as the SVI and CCVI) to predict infection and/or mortality rates but do not always include an application to vaccination uptake [27], [57], [58]. Such studies were included as vulnerability indices, and we note that they are particularly important in comparing the uses of vulnerability indices in order to eventually predict vaccination.

The overlap of utility between the VUIs and vulnerability indices highlighted in our review suggests a potential to refine these tools for improved predictive power. Notably, the use of vulnerability indices (such as the SVI and CCVI) has shown promise in predicting vaccination rates, despite the primary design of such indices for other public health applications [6], [59], [60]. This highlights the potential of existing indices to contribute to next-generation VUIs.

We suggest that integrating both individual- and aggregate-level data alongside temporal and geospatial analyses could enhance the generalizability of these indices. The inclusion of historical vaccination data of other infectious diseases and the application of social and behavioral frameworks could address limitations of current models.

### 4.1 Limitations

First, our search strategy primarily focused on retrieving articles related to COVID-19, given the renewal of interest in vaccination uptake caused by the COVID-19 pandemic. This may have caused an underestimation of the number of vaccine-agnostic studies found to use historical, administrative vaccination data across multiple infectious diseases.

Second, we assumed that vaccine acceptance, hesitancy, and associated factors predict vaccine uptake by extension, but this is an oversimplification [61].

## 5. Conclusions

This scoping review of vaccine uptake indices found that most studies exhibited diverse characteristics of a methodologically advanced vaccine uptake index, though these characteristics were not frequently present together. These studies often utilize geospatial data, but less commonly (1) use individual-level data in tandem with administrative data, (2) integrate geospatial and temporal data, (3) utilize vaccination data sources from multiple infectious diseases, and (4) incorporate social and behavioral frameworks into their study designs. These findings suggest that future research could be performed to develop evidenced-based vaccine uptake indices for the purpose of public health intervention strategies aimed at facilitating equitable vaccination coverage.

## Data Availability

All data produced in the present study are available upon request to the authors.

## Author Contributions

Conceptualization, K.G. and A.G.; Data curation, K.G., S.A., and A.G.; Formal analysis, K.G., S.A., and A.G.; Funding acquisition, K.G., R.H., and A.G.; Investigation, K.G. and S.A.; Methodology, K.G., S.A., R.H., and A.G.; Project administration, K.G.; Resources, K.G.; Software, K.G. and R.H.; Supervision, A.G.; Validation, K.G., S.A., R.H., and A.G.; Visualization, K.G.; Roles/Writing - original draft, K.G. and R.H.; Writing - review & editing, K.G., S.A., R.H., and A.G.

All authors attest they meet the ICMJE criteria for authorship.

## Acknowledgements

Dr. Gutfraind was supported by grant CDC-RFA-DP-19-001 COVID-19 supplemental grant and National Institute of Allergy and Infectious Diseases of the National Institutes of Health (USA) Grant #R01AI158666. We thank the University of Illinois at Chicago Liberal Arts & Sciences Undergraduate Research Initiative. Funding sources had no role in study design; in the collection, analysis and interpretation of data; in the writing of the report; and in the decision to submit the article for publication.

## Supplements

Supplement 1: Search strings used for article retrieval

Supplement 2: Data extraction form

Supplement 3: Further reading of selected articles

Supplement 4: Complete list of articles included by the review

