## Supplement 1 for "Modern approaches to predicting vaccine hesitancy: A scoping review"

**Supplement 1.** Search strings used for article retrieval

| | |
|---|---|
| **Conceptual Domains** | Domain 1: Quantitative Indices<br>Domain 2: Geospatial Analysis<br>Domain 3: Infectious Disease (COVID-19)<br>Domain 4: Vaccination or Vulnerability (Outcomes) |
| **Embase Search String (November 9, 2022)** | ('covid-19 community vulnerability index':ti,ab OR 'social vulnerability index':ti,ab OR 'social vulnerability index'/exp OR 'pandemic vulnerability index':ti,ab OR 'index':ti,ab OR 'indices':ti,ab OR 'quantify':ti,ab OR 'quantitative':ti,ab OR 'model':ti,ab OR 'models':ti,ab OR 'modeling':ti,ab OR 'machine learning'/exp OR 'machine learning':ti,ab OR 'artificial intelligence':ti,ab OR 'algorithm':ti,ab OR 'algorithms':ti,ab OR 'predictor':ti,ab OR 'predictors':ti,ab OR 'predictive':ti,ab OR 'prediction':ti,ab OR 'feature':ti,ab OR 'features':ti,ab OR 'prevention and control'/de) AND ('multi-level':ti,ab OR 'multilevel':ti,ab OR 'census*':ti,ab OR 'census block*':ti,ab OR 'census tract*':ti,ab OR 'mapping':ti,ab OR 'zip code*':ti,ab OR 'county*':ti,ab OR 'community*':ti,ab OR 'neighborhood*':ti,ab OR 'small area':ti,ab OR 'area-based':ti,ab OR 'spatial location':ti,ab OR 'spatial patterns':ti,ab OR 'spatial data':ti,ab OR 'spatial analysis':ti,ab OR 'spatial analysis'/exp OR 'geocod*':ti,ab OR 'geograph*':ti,ab OR 'geospatial':ti,ab OR 'gis':ti,ab OR 'geographic information system'/exp OR 'geographic information system*':ti,ab) AND ('cov':ti,ab OR 'covid':ti,ab OR 'covid19':ti,ab OR 'covid-19':ti,ab OR 'infectious disease*':ti,ab OR 'coronavirus':ti,ab OR 'corona virus':ti,ab OR 'sars-cov-2 vaccine'/de OR 'epidemiology'/de) AND ('vaccin*':ti,ab,kw OR 'immuni*':ti,ab,kw OR 'vulnerab*':ti,ab,kw) |
| **PubMed Search String (November 9, 2022)** | ("COVID-19 community vulnerability index"[tiab] OR "social vulnerability index"[tiab] OR "Social Vulnerability"[mesh] OR "pandemic vulnerability index"[tiab] OR "index"[tiab] OR "indices"[tiab] OR "quantify"[tiab] OR "quantitative"[tiab] OR "model"[tiab] OR "models"[tiab] OR "modeling"[tiab] OR "Machine Learning"[mesh] OR "machine learning"[tiab] OR "artificial intelligence"[tiab] OR "algorithm"[tiab] OR "algorithms"[tiab] OR "predictor"[tiab] OR "predictors"[tiab] OR "predictive"[tiab] OR "prediction"[tiab] OR "feature"[tiab] OR "features"[tiab] OR "Models, Theoretical"[mesh]) AND ("multi-level"[tiab] OR "multilevel"[tiab] OR "census*"[tiab] OR "census block*"[tiab] OR "census tract*"[tiab] OR "mapping"[tiab] OR "zip code*"[tiab] OR |

| | |
|---|---|
| | "county*"[tiab] OR "community*"[tiab] OR "neighborhood*"[tiab] OR "small area"[tiab] OR "Small-area Analysis"[mesh] OR "area-based"[tiab] OR "spatial location"[tiab] OR "spatial patterns"[tiab] OR "spatial data"[tiab] OR "spatial analysis"[tiab] OR "Spatial Analysis"[mesh] OR "geocod*"[tiab] OR "geograph*"[tiab] OR "geospatial"[tiab] OR "GIS"[tiab] OR "Geographic Information Systems"[mesh] OR "geographic information system*"[tiab]) AND ("COVID"[tiab] OR "COVID19"[tiab] OR "COVID-19"[tiab] OR "COVID-19"[mesh] OR "COVID-19 Vaccines"[mesh] OR "infectious disease*"[tiab] OR "coronavirus"[tiab] OR "corona virus"[tiab] OR "coronavirus"[mesh] OR "COV"[tiab] OR "Communicable Diseases"[mesh]) AND ("vaccin*"[tw] OR "immuni*"[tw] OR "vulnerab*"[tw]) |
| **Web of Science Search String (November 7, 2022)** | ("""COVID-19 community vulnerability index"" OR ""social vulnerability index"" OR ""pandemic vulnerability index"" OR ""index"" OR ""indices"" OR ""quantify"" OR ""quantitative"" OR ""model"" OR ""models"" OR ""modeling"" OR ""machine learning"" OR ""artificial intelligence"" OR ""algorithm"" OR ""algorithms"" OR ""predictor"" OR ""predictors"" OR ""predictive"" OR ""prediction"" OR ""feature"" OR ""features""") AND ("""multi-level"" OR ""multilevel"" OR ""census*"" OR ""census block*"" OR ""census tract*"" OR ""mapping"" OR ""zip code*"" OR ""county*"" OR ""community*"" OR ""neighborhood*"" OR ""small area"" OR ""area-based"" OR ""spatial location"" OR ""spatial patterns"" OR ""spatial data"" OR ""spatial analysis"" OR ""geocod*"" OR ""geograph*"" OR ""geospatial"" OR ""GIS"" OR ""geographic information system*""") AND ("""COV"" OR ""COVID"" OR ""COVID19"" OR ""COVID-19"" OR ""infectious disease*"" OR ""coronavirus"" OR ""corona virus""") AND ("""vaccin*"" OR ""immuni*"" OR ""vulnerab*""") |
