## Supplement 2 for "Modern approaches to predicting vaccine hesitancy: A scoping review"

**Supplement 2.** Data extraction form

| | |
|---|---|
| **1. General information** | a) What country or region of the world was this study conducted in? |
| | b) If a) is "United States," what state was the study conducted in? |
| | c) When was the study published (year)? |
| | d) When was the study published (month)? |
| **2. Goal of study** | a) Is the study a vaccine uptake index or a relevant vulnerability index? [Vaccine uptake index, Vulnerability index] |
| **3. Geospatial analysis** | a) Does the study utilize geospatial analysis in its design? |
| | b) If so to a), does the study also utilize temporal analysis in its design? |
| | c) If so to a), what level of geospatial areas were compared? [Multiple select: ZIP code or neighborhood, County or city, State or province, Country] |
| **4. Data types and study design** | a) What type of data was used to create the index? [Multiple select: Individual-level predictors, Aggregate-level predictors] |
| | b) Does the study use other infectious disease data (e.g., historical influenza data) to infer COVID-19 vaccine uptake or vulnerability, or vice-versa? (Does it use vaccine-agnostic methods?) |
| | c) Does the study use a sociobehavioral framework to characterize social vulnerability or vaccination-related behaviors? |
| | d) Please list the sociobehavioral framework if the answer to c) (the previous question) is "yes." |
| | e) Does the study involve the creation of a model that takes geospatial area as an "input" and "outputs" a proportion of the population that is expected to be vaccinated? (Does it adhere to this study's definition of a methodologically advanced vaccine uptake index?) |
