## Supplement 3 for "Modern approaches to predicting vaccine hesitancy: A scoping review"

**Supplement 3.** Further reading of selected articles

| | |
|---|---|
| **Exemplar vaccine uptake and/or novel vulnerability indices** | Avirappattu et al., 2022; Bauer, Zhang, Lee, Fisher-Hoch, et al., 2021; Bauer, Zhang, Lee, Jones, et al., 2021; Bilal et al., 2022; C. Brown et al., 2021; Cheong et al., 2021; Cuadros et al., 2022; DiRago et al., 2022; Filosa et al., 2022; Georges et al., 2022; Hernandez et al., 2022; Jabalameli et al., 2022; Kazemi et al., 2022; Lee et al., 2022; Lee & Huang, 2022; Liao et al., 2022; Liu & Li, 2021; McLaughlin, Khan et al., 2022; M. Mehta et al., 2020; Mirpuri & Rovin, 2021; Mody et al., 2022; Mofleh et al., 2022; Mollalo & Tatar, 2021; Moosazadeh et al., 2022; Tiwari et al., 2021 |
| **Examples of vulnerability indices** | K. Brown et al., 2021; Cartaxo et al., 2021; de Figueiredo et al., 2022; de Souza et al., 2020; Fall et al., 2022; Lee & Ramírez, 2022; Macharia et al., 2020; Madhav et al., 2020; McCoy et al., 2021; M. Mehta et al., 2020; Mofleh et al., 2022; Pathak et al., 2022; Prieto et al., 2021; Relova et al., 2022; Ruck et al., 2021; Saghapour et al., 2021; Sarkar & Chouhan, 2021; Snyder & Parks, 2020; Srivastava et al., 2022; Tiwari et al., 2021; Wiki et al., 2021; Wolkin et al., 2022 |
| **Examples of vaccine hesitancy as a response variable** | Abedin et al., 2021; Adu et al., 2022; Al Awaidy et al., 2022; Al-Mohaithef & Padhi, 2020; Alrajeh et al., 2021; Antwi-Berko et al., 2022; Asmare et al., 2021; Bass et al., 2022; Bennett et al., 2022; Bodas et al., 2023; Buscemi et al., 2023; Cook et al., 2022; Dereje et al., 2022; Ebrahimi et al., 2021; Elsayed et al., 2021; Gaitán-Rossi et al., 2022; Gasteiger et al., 2022; Glampson et al., 2021; Griva et al., 2021; Hossain et al., 2021; Joshi et al., 2022; Kabagenyi et al., 2022; Khubchandani et al., 2021; Kimhi et al., 2022; Morales-García et al., 2022; Mouter et al., 2022; Muhajarine et al., 2021; Oliveira et al., 2021; Omar & Hani, 2021; Orangi et al., 2021; Osur et al., 2022; Ruiz & Bell, 2021; Tan et al., 2022; Tram et al., 2022; Umakanthan et al., 2022; Yu et al., 2022 |
| **Ecological studies involving association with vaccination rates** | Choi et al., 2023; Cuadros et al., 2022; Hernandez et al., 2022; Park, 2022 |
| **Inequalities in vaccination rates within vulnerable groups** | Bagasra et al., 2021; Cheng & Li, 2022; Chevet et al., 2022; Coughenour et al., 2021; Doherty et al., 2021; Gutierrez et al., 2022; Khairat et al., 2022; Klinkhammer et al., 2022; Kusuma & Kant, 2022; Mody et al., 2022; Moore et al., 2021; Myers et al., 2022; Nguyen, Nguyen, et al., 2021; Niño et al., 2021; Rich et al., 2022; Samoa et al., 2022; Siegel et al., 2022; Tsai et al., 2022; Wu, 2022; Yuan et al., 2022 |

| | |
|---|---|
| **Individual psychological factors used to predict vaccine uptake** | Duradoni et al., 2022; Han et al., 2023; Kantor & Kantor, 2021; Liao et al., 2022; Parthasarathi et al., 2022; Reimer et al., 2022; Tagini et al., 2022; Winter et al., 2022 |
| **Social media data used to predict vaccine uptake** | Berning et al., 2022; Guntuku et al., 2021; Jabalameli et al., 2022; Kow et al., 2022; Li et al., 2022; McCarthy et al., 2021; Ogbuokiri et al., 2022; Wilson & Wiysonge, 2020 |
| **Examples of geo-spatiotemporal analysis** | Bauer, Zhang, Lee, Jones, et al., 2021; Gorris et al., 2021; Liu & Li, 2021; Moosazadeh et al., 2022 |
| **Examples of vaccine-agnostic analyses** | Attwell et al., 2021; Barnes & Colagiuri, 2022; Berning et al., 2022; Bonham-Werling et al., 2021; Chevet et al., 2022; Filosa et al., 2022; Gasteiger et al., 2022; Goffe et al., 2021; Lama et al., 2022; Mesele, 2021; Mirpuri & Rovin, 2021; Moreland, Gillezeau, Alpert, et al., 2022; Moreland, Gillezeau, Eugene, et al., 2022; Nair & Wales, 2022; Ruiz & Bell, 2021; Tsai et al., 2022 |
| **Usage of sociobehavioral frameworks** | Al-Hasan et al., 2021; Alrajeh et al., 2021; Asmare et al., 2021; Bhochhibhoya et al., 2021; Chu & Liu, 2021; Ezati Rad et al., 2022; Faturohman et al., 2021; Gao et al., 2022; Goffe et al., 2021; Han et al., 2023; Hayashi et al., 2022; Huynh, Nguyen, Nguyen, & Pham, 2021; Huynh, Nguyen, Nguyen, Lam, et al., 2021; Islam et al., 2021; Jiang et al., 2022; Lama et al., 2022; Lee & You, 2022; Li et al., 2021; Liao et al., 2022; Mangla et al., 2021; S. Mehta et al., 2022; Omar & Hani, 2021; Osur et al., 2022; Park, 2022; Pothisa et al., 2022; Reimer et al., 2022; Ruiz & Bell, 2021; Seangpraw et al., 2022; Urrunaga-Pastor et al., 2021; Wakefield & Khauser, 2021; Zampetakis & Melas, 2021; Zhou et al., 2021 |
