## Supplement 4 for "Modern approaches to predicting vaccine hesitancy: A scoping review"

**Supplement 4.** Complete list of articles included by the review

| Author(s) | Year | Title |
|---|---|---|
| Abedin et al. | 2021 | Willingness to vaccinate against COVID-19 among Bangladeshi adults: Understanding the strategies to optimize vaccination coverage |
| Adu et al. | 2022 | Association between close interpersonal contact and vaccine hesitancy: Findings from a population-based survey in Canada |
| Agaku et al. | 2022 | Geographic, Occupational, and Sociodemographic Variations in Uptake of COVID-19 Booster Doses among Fully Vaccinated US Adults, December 1, 2021, to January 10, 2022 |
| Al Awaidy et al. | 2022 | Knowledge, Attitude, and Acceptability of COVID-19 Vaccine in Oman: A Cross-sectional Study |
| Albatineh et al. | 2022 | Prevalence and factors associated with COVID-19 vaccine acceptance among the general population in Asadabad, Iran: A cross-sectional study |
| AlGethami et al. | 2021 | Awareness and Knowledge Towards Pediatric and Adult COVID-19 Vaccination: A Cross Sectional Community-based Study in Saudi Arabia |
| Al-Hasan et al. | 2021 | Does Seeing What Others Do Through Social Media Influence Vaccine Uptake and Help in the Herd Immunity Through Vaccination? A Cross-Sectional Analysis |
| Alhassan et al. | 2021 | COVID-19 vaccine hesitancy among the adult population in Ghana: Evidence from a pre-vaccination rollout survey |
| Ali et al. | 2022 | Vaccine equity in low and middle income countries: A systematic review and meta-analysis |
| Al-Mohaithef & Padhi | 2020 | Determinants of COVID-19 Vaccine Acceptance in Saudi Arabia: A Web-Based National Survey |
| Alrajeh et al. | 2021 | COVID-19 Vaccine Hesitancy Among the Adult Population in Saudi Arabia |
| Alshahrani et al. | 2022 | Attitude and Willingness to Get COVID-19 Vaccines by a Community Pharmacist in Saudi Arabia: A |

| | | |
|---|---|---|
| | | Cross-Sectional Study |
| Anderson & Ray-Warren | 2022 | Racial-Ethnic Residential Clustering and Early COVID-19 Vaccine Allocations in Five Urban Texas Counties |
| Antwi-Berko et al. | 2022 | Determinants and Variations of COVID-19 Vaccine Uptake and Responses Among Minority Ethnic Groups in Amsterdam, the Netherlands |
| Arling et al. | 2021 | A Data Driven Approach for Prioritizing COVID-19 Vaccinations in the Midwestern United States |
| Asmare et al. | 2021 | Behavioral intention and its predictors toward COVID-19 vaccination among people most at risk of exposure in Ethiopia: Applying the theory of planned behavior model |
| Assoumou et al. | 2022 | Addressing Inequities in SARS-CoV-2 Vaccine Uptake: The Boston Medical Center Health System Experience |
| Attwell et al. | 2021 | Converting the maybes: Crucial for a successful COVID-19 vaccination strategy |
| Avirappattu et al. | 2022 | An optimized machine learning model for identifying socio-economic, demographic and health-related variables associated with low vaccination levels that vary across ZIP codes in California |
| Aygun et al. | 2022 | Aspect Based Twitter Sentiment Analysis on Vaccination and Vaccine Types in COVID-19 Pandemic With Deep Learning |
| Bagasra et al. | 2021 | Racial differences in institutional trust and COVID-19 vaccine hesitancy and refusal |
| Barnes & Colagiuri | 2022 | Drivers of the Intention to Receive a COVID-19 Booster Vaccine: Insights from the UK and Australia |
| Baroroh et al. | 2021 | Willingness to vaccinate against coronavirus disease 2019 and related predictors among non-healthcare personnel in Indonesia |
| Barry et al. | 2021 | Patterns in COVID-19 Vaccination Coverage, by Social Vulnerability and Urbanicity—United States, December 14, 2020-May 1, 2021 |

| | | |
|---|---|---|
| Bass et al. | 2022 | Mapping Perceptual Differences to Understand COVID-19 Beliefs in Those with Vaccine Hesitancy |
| Bauer, Zhang, Lee, Fisher-Hoch, et al. | 2021 | Census Tract Patterns and Contextual Social Determinants of Health Associated With COVID-19 in a Hispanic Population From South Texas: A Spatiotemporal Perspective |
| Bauer, Zhang, Lee, Jones, et al. | 2021 | Real-time geospatial analysis identifies gaps in COVID-19 vaccination in a minority population |
| Bennett et al. | 2022 | Attitudes and personal beliefs about the COVID-19 vaccine among people with COVID-19: A mixed-methods analysis |
| Berning et al. | 2022 | Association of Online Search Trends With Vaccination in the United States: June 2020 Through May 2021 |
| Bhochhibhoya et al. | 2021 | Applying the Health Belief Model for Investigating the Impact of Political Affiliation on COVID-19 Vaccine Uptake |
| Bilal et al. | 2022 | Heterogeneity in Spatial Inequities in COVID-19 Vaccination Across 16 Large US Cities |
| Bodas et al. | 2023 | Overcoming the effect of pandemic fatigue on vaccine hesitancy-Will belief in science triumph? |
| Bogart et al. | 2022 | COVID-19 Vaccine Intentions and Mistrust in a National Sample of Black Americans |
| Bollyky et al. | 2022 | Pandemic preparedness and COVID-19: An exploratory analysis of infection and fatality rates, and contextual factors associated with preparedness in 177 countries, from Jan 1, 2020, to Sept 30, 2021 |
| Bonham-Werling et al. | 2021 | Using Statewide Electronic Health Record and Influenza Vaccination Data to Plan and Prioritize COVID-19 Vaccine Outreach and Communications in Wisconsin Communities |
| C. Brown et al. | 2021 | COVID-19 vaccination rates vary by community vulnerability: A county-level analysis |
| K. Brown et al. | 2021 | An ecological study of the association between neighborhood racial and economic residential segregation with COVID-19 vulnerability in the United |

| | | |
|---|---|---|
| | | States' capital city |
| Buscemi et al. | 2023 | Factors Associated With COVID-19 Vaccination Uptake in Community Health Center Patients |
| Cartaxo et al. | 2021 | The exposure risk to COVID-19 in most affected countries: A vulnerability assessment model |
| Carter et al. | 2022 | COVID-19 Vaccine Uptake in Southeastern Ontario, Canada: Monitoring and Addressing Health Inequities |
| Cerio et al. | 2021 | Relationship Between COVID-19 Cases and Vaccination Rates in New York State Counties |
| Cheng & Li | 2022 | Racial and ethnic and income disparities in COVID-19 vaccination among Medicare beneficiaries |
| Cheong et al. | 2021 | Predictive modeling of vaccination uptake in US counties: A machine learning-based approach |
| Chevet et al. | 2022 | COVID-19 Vaccine Uptake Among Patients With Systemic Lupus Erythematosus in the American Midwest: The Lupus Midwest Network (LUMEN) |
| Choi et al. | 2023 | The US COVID-19 surveillance environment: An ecological analysis of the relationship of testing adequacy in the context of vaccination |
| Chu & Liu | 2021 | Integrating health behavior theories to predict American's intention to receive a COVID-19 vaccine |
| Cook et al. | 2022 | Vaccination against COVID-19: Factors That Influence Vaccine Hesitancy among an Ethnically Diverse Community in the UK |
| Coughenour et al. | 2021 | Assessing Determinants of COVID-19 Vaccine Hesitancy in Nevada |
| Cuadros et al. | 2022 | Association Between Vaccination Coverage Disparity and the Dynamics of the COVID-19 Delta and Omicron Waves in the US |
| de Figueiredo et al. | 2022 | Measuring COVID-19 Vulnerability for Northeast Brazilian Municipalities: Social, Economic, and Demographic Factors Based on Multiple Criteria and Spatial Analysis |
| de Souza et al. | 2020 | Human development, social vulnerability and COVID-19 in |

| | | |
|---|---|---|
| | | Brazil: A study of the social determinants of health |
| Dereje et al. | 2022 | COVID-19 vaccine hesitancy in Addis Ababa, Ethiopia: A mixed-method study |
| Diesel et al. | 2021 | COVID-19 Vaccination Coverage Among Adults—United States, December 14, 2020-May 22, 2021 |
| DiRago et al. | 2022 | COVID-19 Vaccine Rollouts and the Reproduction of Urban Spatial Inequality: Disparities Within Large US Cities in March and April 2021 by Racial/Ethnic and Socioeconomic Composition |
| Ditekemena et al. | 2021 | COVID-19 Vaccine Acceptance in the Democratic Republic of Congo: A Cross-Sectional Survey |
| Doherty et al. | 2021 | COVID-19 vaccine hesitancy in underserved communities of North Carolina |
| Duradoni et al. | 2022 | Italian version of the Vaccination Fear Scale (VFS-6): Internal and external validation |
| Ebrahimi et al. | 2021 | Risk, Trust, and Flawed Assumptions: Vaccine Hesitancy During the COVID-19 Pandemic |
| Elsayed et al. | 2021 | Factors Influencing Decision Making Regarding the Acceptance of the COVID-19 Vaccination in Egypt: A Cross-Sectional Study in an Urban, Well-Educated Sample |
| Ezati Rad et al. | 2022 | Predicting the COVID-19 vaccine receive intention based on the theory of reasoned action in the south of Iran |
| Faes et al. | 2022 | Geographical Variation of COVID-19 Vaccination Coverage, Ethnic Diversity and Population Composition in Flanders |
| Fall et al. | 2022 | County-Level Assessment of Vulnerability to COVID-19 in Alabama |
| Faturohman et al. | 2021 | Factors influencing COVID-19 vaccine acceptance in Indonesia: An adoption of Technology Acceptance Model |
| Ferwana & Varshney | 2021 | Social capital dimensions are differentially associated with COVID-19 vaccinations, masks, and physical distancing |

| | | |
|---|---|---|
| Filosa et al. | 2022 | COVID-19 needs no passport: The interrelationship of the COVID-19 pandemic along the U.S.-Mexico border |
| Franic | 2022 | What Lies Behind Substantial Differences in COVID-19 Vaccination Rates Between EU Member States? |
| Gaitán-Rossi et al. | 2022 | Barriers to COVID-19 vaccination among older adults in Mexico City |
| Gao et al. | 2022 | Associations between Public Fear of COVID-19 and Number of COVID-19 Vaccinations: A County-Level Longitudinal Analysis |
| Gasteiger et al. | 2022 | Characteristics associated with the willingness to receive a COVID-19 vaccine and an exploration of the general public's perceptions: A mixed-methods approach |
| Gaughan et al. | 2023 | COVID-19 vaccination uptake amongst ethnic minority communities in England: A linked study exploring the drivers of differential vaccination rates |
| Georges et al. | 2022 | Community-level characteristics of COVID-19 vaccine hesitancy in England: A nationwide cross-sectional study |
| Gerretsen et al. | 2021 | Vaccine Hesitancy Is a Barrier to Achieving Equitable Herd Immunity Among Racial Minorities |
| Glampson et al. | 2021 | Assessing COVID-19 Vaccine Uptake and Effectiveness Through the North West London Vaccination Program: Retrospective Cohort Study |
| Goffe et al. | 2021 | Factors associated with vaccine intention in adults living in England who either did not want or had not yet decided to be vaccinated against COVID-19 |
| Gorris et al. | 2021 | A time-varying vulnerability index for COVID-19 in New Mexico, USA using generalized propensity scores |
| Griva et al. | 2021 | Evaluating Rates and Determinants of COVID-19 Vaccine Hesitancy for Adults and Children in the Singapore Population: Strengthening Our Community's Resilience against Threats from Emerging Infections (SOCRATEs) Cohort |

| | | |
|---|---|---|
| Guntuku et al. | 2021 | Twitter discourse reveals geographical and temporal variation in concerns about COVID-19 vaccines in the United States |
| Gutierrez et al. | 2022 | Predictors of COVID-19 Vaccination Likelihood Among Reproductive-Aged Women in the United States |
| Han et al. | 2023 | Why some people do not get vaccinated against COVID-19: Social-cognitive determinants of vaccination behavior |
| Harapan et al. | 2020 | Acceptance of a COVID-19 Vaccine in Southeast Asia: A Cross-Sectional Study in Indonesia |
| Harris et al. | 2023 | Communities with an anchor institution have higher coronavirus vaccination rates |
| Hayashi et al. | 2022 | Predicting Intention to Take a COVID-19 Vaccine in the United States: Application and Extension of Theory of Planned Behavior |
| Hernandez et al. | 2022 | Disparities in distribution of COVID-19 vaccines across US counties: A geographic information system–based cross-sectional study |
| Herrera-Añazco et al. | 2021 | Prevalence and factors associated with the intention to be vaccinated against COVID-19 in Peru |
| Hossain et al. | 2021 | COVID-19 vaccine hesitancy among the adult population in Bangladesh: A nationwide cross-sectional survey |
| Hu et al. | 2022 | COVID-19 vaccine hesitancy cannot fully explain disparities in vaccination coverage across the contiguous United States |
| Hughes et al. | 2021 | County-Level COVID-19 Vaccination Coverage and Social Vulnerability—United States, December 14, 2020-March 1, 2021 |
| Huynh, Nguyen, Nguyen, & Pham | 2021 | Development and Psychometric Properties of the Health Belief Scales Toward COVID-19 Vaccine in Ho Chi Minh City, Vietnam |
| Huynh, Nguyen, Nguyen, Lam, et al. | 2021 | Knowledge About COVID-19, Beliefs and Vaccination Acceptance Against COVID-19 Among High-Risk People in Ho Chi Minh City, Vietnam |
| Islam et al. | 2021 | Knowledge, attitudes and perceptions |

| | | |
|---|---|---|
| | | towards COVID-19 vaccinations: A cross-sectional community survey in Bangladesh |
| Jabalameli et al. | 2022 | Spatial and sentiment analysis of public opinion toward COVID-19 pandemic using twitter data: At the early stage of vaccination |
| Jiang et al. | 2022 | Changes of COVID-19 Knowledge, Attitudes, Practices and Vaccination Willingness Among Residents in Jinan, China |
| Joshi et al. | 2022 | A cross sectional study to examine factors influencing COVID-19 vaccine acceptance, hesitancy and refusal in urban and rural settings in Tamil Nadu, India |
| Kabagenyi et al. | 2022 | Factors Associated with COVID-19 Vaccine Hesitancy in Uganda: A Population-Based Cross-Sectional Survey |
| Kalayou & Awol | 2022 | Myth and Misinformation on COVID-19 Vaccine: The Possible Impact on Vaccination Refusal Among People of Northeast Ethiopia: A Community-Based Research |
| Kantor & Kantor | 2021 | Development and validation of the Oxford Pandemic Attitude Scale-COVID-19 (OPAS-C): An internet-based cross-sectional study in the UK and USA |
| Kazemi et al. | 2022 | Assessing Inequities in COVID-19 Vaccine Roll-Out Strategy Programs: A Cross-Country Study Using a Machine Learning Approach |
| Khairat et al. | 2022 | Factors and reasons associated with low COVID-19 vaccine uptake among highly hesitant communities in the US |
| Khubchandani et al. | 2021 | COVID-19 Vaccination Hesitancy in the United States: A Rapid National Assessment |
| Kimhi et al. | 2022 | Impact of societal resilience on vaccine hesitancy and uptake: Lessons learned from the Israeli experience |
| Kipnis et al. | 2021 | Evaluation of Vaccination Strategies to Compare Efficient and Equitable Vaccine Allocation by Race and Ethnicity Across Time |
| Klinkhammer et al. | 2022 | Sociopolitical, mental health, and sociodemographic correlates of COVID-19 vaccine hesitancy among |

| | | |
|---|---|---|
| | | young adults in 6 US metropolitan areas |
| Kourlaba et al. | 2021 | Willingness of Greek general population to get a COVID-19 vaccine |
| Kow et al. | 2022 | COVID-19 Infodemiology: Association Between Google Search and Vaccination in Malaysian Population |
| Kusuma & Kant | 2022 | COVID-19 vaccine acceptance and its determinants: A cross-sectional study among the socioeconomically disadvantaged communities living in Delhi, India |
| Lama et al. | 2022 | Factors Associated With COVID-19 Behavioral Intentions: Findings From an Online Survey |
| Lee et al. | 2022 | Epidemic Vulnerability Index for Effective Vaccine Distribution Against Pandemic |
| Lee & Huang | 2022 | COVID-19 Vaccine Hesitancy: The Role of Socioeconomic Factors and Spatial Effects |
| Lee & Ramírez | 2022 | Geography of Disparity: Connecting COVID-19 Vulnerability and Social Determinants of Health in Colorado |
| Lee & You | 2022 | Direct and Indirect Associations of Media Use With COVID-19 Vaccine Hesitancy in South Korea: Cross-sectional Web-Based Survey |
| Lu Li et al. | 2021 | The intention to receive the COVID-19 vaccine in China: Insights from protection motivation theory |
| Lingyao Li et al. | 2022 | Dynamic assessment of the COVID-19 vaccine acceptance leveraging social media data |
| Liao | 2021 | Social and economic inequality in coronavirus disease 2019 vaccination coverage across Illinois counties |
| Liao et al. | 2022 | Priming with social benefit information of vaccination to increase acceptance of COVID-19 vaccines |
| Lindholt et al. | 2021 | Public acceptance of COVID-19 vaccines: Cross-national evidence on levels and individual-level predictors using observational data |
| Liu & Li | 2021 | Hesitancy in the time of coronavirus: Temporal, spatial, and sociodemographic variations in COVID-19 vaccine hesitancy |

| | | |
|---|---|---|
| Macharia et al. | 2020 | A vulnerability index for COVID-19: Spatial analysis at the subnational level in Kenya |
| Madhav et al. | 2020 | The effect of area deprivation on COVID-19 risk in Louisiana |
| Mangla et al. | 2021 | COVID-19 Vaccine Hesitancy and Emerging Variants: Evidence from Six Countries |
| Martinez et al. | 2022 | Equitable COVID-19 Vaccination for Hispanics in the United States: A Success Story from California Border Communities |
| Marvel et al. | 2021 | The COVID-19 Pandemic Vulnerability Index (PVI) Dashboard: Monitoring county-level vulnerability using visualization, statistical modeling, and machine learning |
| Maugeri et al. | 2022 | Using Google Trends to Predict COVID-19 Vaccinations and Monitor Search Behaviours about Vaccines: A Retrospective Analysis of Italian Data |
| McCarthy et al. | 2021 | Social Data: An Underutilized Metric for Determining Participation in COVID-19 Vaccinations |
| McCoy et al. | 2021 | Ensemble machine learning of factors influencing COVID-19 across US counties |
| McLaughlin, Khan, et al. | 2022 | County-level vaccination coverage and rates of COVID-19 cases and deaths in the United States: An ecological analysis |
| McLaughlin, Wiemken, et al. | 2022 | US County-Level COVID-19 Vaccine Uptake and Rates of Omicron Cases and Deaths |
| M. Mehta et al. | 2020 | Early Stage Machine Learning-Based Prediction of US County Vulnerability to the COVID-19 Pandemic: Machine Learning Approach |
| S. Mehta et al. | 2022 | Knowledge, Attitude, Practices, and Vaccine Hesitancy Among the Latinx Community in Southern California Early in the COVID-19 Pandemic: Cross-sectional Survey |
| Mesele | 2021 | COVID-19 Vaccination Acceptance and Its Associated Factors in Sodo Town, Wolaita Zone, Southern Ethiopia: Cross-Sectional Study |
| Minaya et al. | 2022 | Medical Mistrust, COVID-19 Stress, |

| | | |
|---|---|---|
| | | and Intent to Vaccinate in Racial-Ethnic Minorities |
| Mirpuri & Rovin | 2021 | COVID-19 and Historic Influenza Vaccinations in the United States: A Comparative Analysis |
| Mody et al. | 2022 | Quantifying inequities in COVID-19 vaccine distribution over time by social vulnerability, race and ethnicity, and location: A population level analysis in St. Louis and Kansas City, Missouri |
| Mofleh et al. | 2022 | Spatial Patterns of COVID-19 Vaccination Coverage by Social Vulnerability Index and Designated COVID-19 Vaccine Sites in Texas |
| Mollalo & Tatar | 2021 | Spatial Modeling of COVID-19 Vaccine Hesitancy in the United States |
| Moola et al. | 2021 | A rapid review of evidence on the determinants of and strategies for COVID-19 vaccine acceptance in low- and middle-income countries |
| Moore et al. | 2021 | Correlates of COVID-19 Vaccine Hesitancy among a Community Sample of African Americans Living in the Southern United States |
| Moosazadeh et al. | 2022 | A machine learning-driven spatio-temporal vulnerability appraisal based on socio-economic data for COVID-19 impact prevention in the US counties |
| Morales-García et al. | 2022 | Predictors of Intention to Vaccinate Against COVID-19 in a Peruvian Sample |
| Moreland, Gillezeau, Alpert, et al. | 2022 | Assessing influenza vaccination success to inform COVID-19 vaccination campaign |
| Moreland, Gillezeau, Eugene, et al. | 2022 | Ecologic study of influenza vaccination uptake and COVID-19 death rate in New York City |
| Mouter et al. | 2022 | "Please, you go first!" preferences for a COVID-19 vaccine among adults in the Netherlands |
| Muchiri et al. | 2022 | Unmet need for COVID-19 vaccination coverage in Kenya |
| Muhajarine et al. | 2021 | COVID-19 vaccine hesitancy and refusal and associated factors in an adult population in Saskatchewan, Canada: Evidence from predictive |

| | | |
|---|---|---|
| | | modelling |
| Murthy et al. | 2021 | Disparities in COVID-19 Vaccination Coverage Between Urban and Rural Counties—United States, December 14, 2020-April 10, 2021 |
| Mushtaq et al. | 2022 | Analyses of Public Attention and Sentiments towards Different COVID-19 Vaccines Using Data Mining Techniques |
| Myers et al. | 2022 | COVID-19 vaccination hesitancy among Americans with disabilities aged 18-65: An exploratory analysis |
| Nafilyan et al. | 2021 | Sociodemographic inequality in COVID-19 vaccination coverage among elderly adults in England: A national linked data study |
| Nair & Wales | 2022 | Seasonal and 2009 Pandemic H1N1 Vaccine Acceptance as a Predictor for COVID-19 Vaccine Acceptance |
| Nguyen, Anneser, et al. | 2022 | Disparities in national and state estimates of COVID-19 vaccination receipt and intent to vaccinate by race/ethnicity, income, and age group among adults ≥ 18 years, United States |
| Nguyen, Nguyen, et al. | 2021 | Changes in COVID-19 vaccination receipt and intention to vaccinate by socioeconomic characteristics and geographic area, United States, January 6–March 29, 2021 |
| Niño et al. | 2021 | Trajectories of COVID-19 vaccine intentions among U.S. adults: The role of race and ethnicity |
| Ogbuokiri et al. | 2022 | Public sentiments toward COVID-19 vaccines in South African cities: An analysis of Twitter posts |
| Oliveira et al. | 2021 | Prevalence and factors associated with covid-19 vaccine hesitancy in Maranhão, Brazil |
| Omar & Hani | 2021 | Attitudes and intentions towards COVID-19 vaccines and associated factors among Egyptian adults |
| Omidvar Tehrani & Perkins | 2022 | Community Health Resources, Globalization, Trust in Science, and Voting as Predictors of COVID-19 Vaccination Rates: A Global Study with Implications for Vaccine Adherence |
| Ong et al. | 2021 | COVID-19 Medical Vulnerability |

| | | |
|---|---|---|
| | | Indicators: A Predictive, Local Data Model for Equity in Public Health Decision Making |
| Orangi et al. | 2021 | Assessing the Level and Determinants of COVID-19 Vaccine Confidence in Kenya |
| Osur et al. | 2022 | Determinants of COVID-19 vaccine behaviour intentions among the youth in Kenya: A cross-sectional study |
| Ozdenerol & Seboly | 2022 | The Effects of Lifestyle on COVID-19 Vaccine Hesitancy in the United States: An Analysis of Market Segmentation |
| Pan et al. | 2022 | Factors That Impact Acceptance of COVID-19 Vaccination in Different Community-Dwelling Populations in China |
| Park | 2022 | Regional Disparities in COVID-19 Vaccine Hesitancy: The Moderating Role of Social Distancing and Vaccine Rollout in the US |
| Parthasarathi et al. | 2022 | Willingness to Accept the COVID-19 Vaccine and Related Factors among Indian Adults: A Cross-Sectional Study |
| Pathak et al. | 2022 | Assessing Socioeconomic Vulnerabilities Related to COVID-19 Risk in India: A State-Level Analysis |
| Paudel et al. | 2022 | The influence of place on COVID-19 vaccine coverage in Alberta: A multilevel analysis |
| Perry et al. | 2021 | Inequalities in coverage of COVID-19 vaccination: A population register based cross-sectional study in Wales, UK |
| Porteny et al. | 2022 | Associations among political voting preference, high-risk health status, and preventative behaviors for COVID-19 |
| Pothisa et al. | 2022 | Knowledge of COVID-19 and Its Relationship with Preventive Behaviors and Vaccination among Adults in Northern Thailand’s Community |
| Prieto et al. | 2021 | Urban Vulnerability Assessment for Pandemic Surveillance-The COVID-19 Case in Bogota, Colombia |
| Putri et al. | 2022 | Acceptance of Covid 19 Vaccine in Terms of Perception and Knowledge: A Cross-Sectional Study |

| | | |
|---|---|---|
| Qiao et al. | 2022 | Social Capital, Urbanization Level, and COVID-19 Vaccination Uptake in the United States: A National Level Analysis |
| Reimer et al. | 2022 | Moral values predict county-level COVID-19 vaccination rates in the United States |
| Relova et al. | 2022 | British Columbia’s Index of Multiple Deprivation for Community Health Service Areas |
| Renzi et al. | 2022 | Mapping the Prevalence of COVID-19 Vaccine Acceptance at the Global and Regional Level: A Systematic Review and Meta-Analysis |
| Rich et al. | 2022 | How education and racial segregation intersect in neighborhoods with persistently low COVID-19 vaccination rates in Philadelphia |
| Rovetta | 2022 | Google Trends as a Predictive Tool for COVID-19 Vaccinations in Italy: Retrospective Infodemiological Analysis |
| Roy et al. | 2022 | Factors influencing COVID-19 vaccine acceptance and hesitancy among rural community in Bangladesh: A cross-sectional survey based study |
| Ruck et al. | 2021 | Early warning of vulnerable counties in a pandemic using socio-economic variables |
| Ruiz & Bell | 2021 | Predictors of intention to vaccinate against COVID-19: Results of a nationwide survey |
| Ryerson et al. | 2021 | Disparities in COVID-19 Vaccination Status, Intent, and Perceived Access for Noninstitutionalized Adults, by Disability Status—National Immunization Survey Adult COVID Module, United States, May 30-June 26, 2021 |
| Saghapour et al. | 2021 | Supporting pandemic disease preparedness: Development of a composite index of area vulnerability |
| Samoa et al. | 2022 | Socioeconomic Inequities in Vaccine Hesitancy Among Native Hawaiians and Pacific Islanders |
| Sarkar & Chouhan | 2021 | COVID-19: District level vulnerability assessment in India |
| Sarkar et al. | 2022 | COVID-19 Vulnerability Mapping of |

| | | |
|---|---|---|
| | | Asian Countries |
| Seale et al. | 2021 | Examining Australian public perceptions and behaviors towards a future COVID-19 vaccine |
| Seangpraw et al. | 2022 | Using the Health Belief Model to Predict Vaccination Intention Among COVID-19 Unvaccinated People in Thai Communities |
| Sethi et al. | 2021 | The UPTAKE study: A cross-sectional survey examining the insights and beliefs of the UK population on COVID-19 vaccine uptake and hesitancy |
| Siegel et al. | 2022 | Racial/Ethnic Disparities in State-Level COVID-19 Vaccination Rates and Their Association with Structural Racism |
| Snyder & Parks | 2020 | Spatial variation in socio-ecological vulnerability to Covid-19 in the contiguous United States |
| Sokale et al. | 2022 | COVID-19 Vaccine Uptake among US Adults According to Standard Occupational Groups |
| Sun & Monnat | 2022 | Rural-urban and within-rural differences in COVID-19 vaccination rates |
| Sun & Rhubart | 2022 | Rural-Urban Differences in the Associations Between Aging and Disability Services and COVID-19 Vaccination Rates Among Older Adults |
| Tagini et al. | 2022 | Behind the Scenes of COVID-19 Vaccine Hesitancy: Psychological Predictors in an Italian Community Sample |
| Tan et al. | 2022 | Determining the Prevalence and Correlates of COVID-19 Booster Vaccine Hesitancy in the Singapore Population Following the Completion of the Primary Vaccination Series |
| Thakore et al. | 2021 | Association of Social Vulnerability, COVID-19 vaccine site density, and vaccination rates in the United States |
| Thompson et al. | 2021 | Factors Associated with Racial/Ethnic Group-Based Medical Mistrust and Perspectives on COVID-19 Vaccine Trial Participation and Vaccine Uptake in the US |

| | | |
|---|---|---|
| Tipirneni et al. | 2022 | Associations of 4 Geographic Social Vulnerability Indices With US COVID-19 Incidence and Mortality |
| Tiu et al. | 2022 | Characterizing the Spatiotemporal Heterogeneity of the COVID-19 Vaccination Landscape |
| Tiwari et al. | 2021 | Using machine learning to develop a novel COVID-19 Vulnerability Index (C19VI) |
| Tortolero et al. | 2022 | Examining Social Vulnerability and the Association With COVID-19 Incidence in Harris County, Texas |
| Tram et al. | 2022 | Deliberation, Dissent, and Distrust: Understanding Distinct Drivers of Coronavirus Disease 2019 Vaccine Hesitancy in the United States |
| Travis et al. | 2021 | Identifying the determinants of COVID-19 preventative behaviors and vaccine intentions among South Carolina residents |
| Tsai et al. | 2022 | COVID-19 Vaccine Hesitancy and Acceptance Among Individuals With Cancer, Autoimmune Diseases, or Other Serious Comorbid Conditions: Cross-sectional, Internet-Based Survey |
| Umakanthan et al. | 2022 | Social Environmental Predictors of COVID-19 Vaccine Hesitancy in India: A Population-Based Survey |
| Urrunaga-Pastor et al. | 2021 | Cross-sectional analysis of COVID-19 vaccine intention, perceptions and hesitancy across Latin America and the Caribbean |
| Valckx et al. | 2022 | Individual factors influencing COVID-19 vaccine acceptance in between and during pandemic waves (July–December 2020) |
| Wagner et al. | 2022 | Vaccine Hesitancy During the COVID-19 Pandemic: A Latent Class Analysis of Middle-Aged and Older US Adults |
| Wakefield & Khauser | 2021 | Doing it for us: Community identification predicts willingness to receive a COVID-19 vaccination via perceived sense of duty to the community |
| J. Wang et al. | 2022 | From COVID-19 Vaccination Intention to Actual Vaccine Uptake: A Longitudinal Study Among Chinese Adults After Six Months of a National |

| | | |
|---|---|---|
| | | Vaccination Campaign |
| Q. Wang et al. | 2022 | Governmental Incentives, Satisfaction with Health Promotional Materials, and COVID-19 Vaccination Uptake among Community-Dwelling Older Adults in Hong Kong: A Random Telephone Survey |
| Z. Wang et al. | 2022 | Mapping global acceptance and uptake of COVID-19 vaccination: A systematic review and meta-analysis |
| Welsh et al. | 2022 | Static Socio-Ecological COVID-19 Vulnerability Index and Vaccine Hesitancy Index for England |
| Weston et al. | 2022 | Targeting Equity in COVID-19 Vaccinations Using the “Evaluating Vulnerability and Equity” (EVE) Model |
| Whiteman et al. | 2021 | Demographic and Social Factors Associated with COVID-19 Vaccination Initiation Among Adults Aged ≥65 Years—United States, December 14, 2020-April 10, 2021 |
| Wiki et al. | 2021 | Understanding vulnerability to COVID-19 in New Zealand: A nationwide cross-sectional study |
| Wilson et al. | 2021 | Health Intelligence Atlas: A Core Tool for Public Health Intelligence |
| Wilson & Wiysonge | 2020 | Social media and vaccine hesitancy |
| Winter et al. | 2022 | Conspiracy beliefs and distrust of science predicts reluctance of vaccine uptake of politically right-wing citizens |
| Wojcicki et al. | 2022 | Household and social characteristics associated with COVID-19 vaccine intent among Latino families in the San Francisco Bay Area |
| Wolkin et al. | 2022 | Comparison of National Vulnerability Indices Used by the Centers for Disease Control and Prevention for the COVID-19 Response |
| Wu | 2022 | Racial concentration and dynamics of COVID-19 vaccination in the United States |
| J. Wu et al. | 2022 | COVID-19 Vaccination Acceptance Among Chinese Population and Its Implications for the Pandemic: A National Cross-Sectional Study |
| T.-Y. Wu et al. | 2022 | Perceptions of COVID-19 Vaccine, Racism, and Social Vulnerability: An |

| | | |
|---|---|---|
| | | Examination among East Asian Americans, Southeast Asian Americans, South Asian Americans, and Others |
| Xu & Jiang | 2022 | Effects of Social Vulnerability and Spatial Accessibility on COVID-19 Vaccination Coverage: A Census-Tract Level Study in Milwaukee County, USA |
| Yu et al. | 2022 | COVID-19 vaccine hesitancy and resistance in an urban Chinese population of Hong Kong: A cross-sectional study |
| Yuan et al. | 2022 | Race, ethnicity, psychological factors, and COVID-19 vaccine hesitancy during the COVID-19 pandemic |
| Zampetakis et al. | 2021 | The health belief model predicts vaccination intentions against COVID-19: A survey experiment approach |
| Zeng et al. | 2022 | Association of Zip Code Vaccination Rate With COVID-19 Mortality in Chicago, Illinois |
| Zhong et al. | 2022 | Sparse spatially clustered coefficient model via adaptive regularization |
| Zhou et al. | 2021 | COVID-19 vaccination acceptance in china after it becomes available: A cross-sectional study |
